# A comparison of different methods for handling measurements affected by medication use

**DOI:** 10.1101/2022.04.23.22273899

**Authors:** Jungyeon Choi, Olaf M. Dekkers, Saskia le Cessie

**Affiliations:** Department of Clinical Epidemiology, Leiden University Medical Center, Address: Albinusdreef 2, C7-P, 2333 ZA Leiden, the Netherlands; Department of Clinical Epidemiology & Department of Endocrinology and Metabolism, Leiden, University Medical Center, Address: Albinusdreef 2, C7-P, 2333 ZA Leiden, the Netherlands; Department of Clinical Epidemiology & Department of Biomedical Data sciences, Leiden University, Medical Center, Address: Albinusdreef 2, C7-P, 2333 ZA Leiden, the Netherlands

**Author notes:** Corresponding author: Jungyeon Choi. Declarations of interest: none.

**Keywords:** Medication effect, measurement error, censored data, missing data, regression calibration, multiple imputation

## Abstract

In epidemiological research it is common to encounter measurements affected by medication use, such as blood pressure lowered by antihypertensive drugs. When one is interested in the relation between the variables not affected by medication, ignoring medication use can cause bias. Several methods have been proposed, but the problem is often ignored or handled with generic methods, such as excluding individuals on medication or adjusting for medication use in the analysis. This study aimed to investigate methods for handling measurements affected by medication use when one is interested in the relation between the unaffected variables and to provide guidance for how to optimally handle the problem. We focused on linear regression and distinguish between the situation where the affected measurement is an exposure, confounder or outcome. In the Netherlands Epidemiology of Obesity study and in several simulated settings, we compared generic and more advanced methods, such as substituting or adding a fixed value to the treated values, regression calibration, censored normal regression, Heckman’s treatment model and multiple imputation methods. We found that often-used methods such as adjusting for medication use could result in substantial bias and that methods for handling medication use should be chosen cautiously.

## 1. Introduction

Measurements affected by medication use are commonly encountered in epidemiological research. Examples are glucose levels lowered by glucose lowering medications or blood pressure relieved by antihypertensive drugs. Depending on research questions, these measurements can be an outcome of interest or covariates.

Although researchers often are interested in the effect of certain drugs, the relation between the values not affected by medication can also be the primary scientific interest. However, the value of a variable had an individual not been treated is often not available. Using the values affected by medication instead may lead to biased results. In clinical research, however, medication use is often ignored or handled with naïve methods such as excluding medication users or adjusting for medication use. For outcomes affected by medication use, these naïve methods may introduce bias (1-4).

Several methods have been proposed to handle measurements affected by medication use. Relatively simple methods are adding an expected medication effect to treated values or substituting the treated values for other values (1, 4, 5). More sophisticated methods include censored normal regression, Heckman’s treatment model, quantile regression, measurement error methods or advanced imputation techniques (1, 2, 6, 7). However, these methods are seldom used in applied research. Additionally, many of the suggested methods are limited to outcomes affected by medication, and little has been known on how to handle exposures or confounders affected by medication.

The aim of this study is to investigate methods for handling measurements affected by medication use when the unaffected values are of interest. We focused on etiological studies where effects are estimated by linear regression. We discuss different methods and compare these methods in a large cross-sectional study of the Netherlands Epidemiology of Obesity (NEO) study and in several simulation scenarios generated based on the NEO data. The scenarios vary on whether the exposure, confounder or the outcome is affected by medication use. Based on the results of simulation study, we provide guidance on how to optimally handle the medication effect.

## 2. Methods to handle measurements affected by medication use

We will consider the situation where for some individuals a variable is affected by medication use (e.g. blood pressure affected by antihypertensive drug), while the relation between variables when no one is affected by medication is of interest. For convenience, we assume that medication is taken when values are high, aiming to lower the values. Depending on the research question, the variable affected by medication use can be the exposure, a confounder or the outcome in an analysis.

The problem of measurements affected by medication can be viewed from different perspectives; it can be viewed as a missing data problem, because for people on medication their untreated values are unobserved. It may be viewed as a measurement error problem, as the observed values differ systematically from the values had the treated individuals not been treated. It could also be viewed as a censoring problem, if we assume that the unobserved untreated values are at least as high as the observed values under treatment. Depending on how one approaches the problem, methods for missing data, for measurement error or for censored observations can be used.

Table 1 summarizes methods for handling measurements affected by medication use. The methods can be categorized as generic methods [M1-M5], a method for the exposure affected by medication [M6], methods for the outcome affected by medication [M7-M10], and multiple imputation approaches [M11-M13]. Detailed descriptions on each method and underlying assumptions are available in Supplementary Material 1. All methods are applied to empirical and simulated data in the following sections.

**Table 1.** Overview of Methods for Handling Measurements Affected by Medication use.

|  | Methods | Description |
| --- | --- | --- |
| Generic methods | **[M1]** Ignoring medication use | Medication use is ignored. |
|  | **[M2]** Restricting to untreated individuals | The analysis is performed in the subgroup of individuals who are not receiving medication. |
|  | **[M3]** Binary adjustment for medication use | An indicator for medication use is added as a covariate in the regression model. |
|  | **[M4]** Substituting measurement of treated individuals with a fixed value | Measurements affected by medication are substituted with a pre-specified value. |
|  | **[M5]** Adding a constant value to observations of treated individuals | A prespecified treatment effect is added to the observed measurements of treated individuals. |
| For exposures affected by medication | **[M6]** Regression calibration | Measurement error methods are used. Based on the expected mean treatment effect and its standard deviation, the observed measurements affected by medication are corrected . |
| For outcomes affected by medication | **[M7]** Inverse probability weighting | Treated individuals are removed in the analysis, and a reweighted analysis is performed where more weight is given to individuals who are untreated but have similar profile as treated individuals |
|  | **[M8]** Quantile regression | The method assumes that for the untreated values for individuals on medication would have been above the median, conditional on covariates. The median outcome is modelled as function of covariates. |
|  | **[M9]** Censored normal regression | Measurements of treated individuals are considered to be censored observations, where the untreated values are assumed to be at least as high as the observed vales affected by treatment, or in more complex censoring mechanisms, at least as high as the observed values and a clinical guideline at which treatment is prescribed. |
|  | **[M10]** Heckman's treatment model | Treatment assignment is assumed to be dependent on the untreated values, and the treatment results in a “structural shift” of the mean outcome. |
| Multiple imputation approaches | **[M11]** Predictive mean matching | A default multiple imputation option in commonly used statistical software. It assumes that the observations of treated individuals are missing at random. |
|  | **[M12]** Censored normal imputation | Censored normal regression is used in the imputation algorithm to predict the untreated values of those on treatment |
|  | **[M13]** Heckman's model imputation | Heckman's model is used in the imputation algorithm to predict the untreated values of those on treatment |

## 3. Example: the Netherlands Epidemiology of Obesity Study

The Netherlands Epidemiology of Obesity (NEO) study is a population-based study designed to investigate pathways that lead to obesity-related diseases. From 2008 to 2012, 6,671 individuals aged 45–65 years were included in the study. Participants brought all medication they were using to the NEO study site, which was coded using the Anatomical Therapeutic Chemical Classification (8). Details can be found elsewhere (9). The NEO study data includes several measurements affected by medication, for example, 31% of the participants used antihypertensive medication and 15% used lipid lowering medication.

To illustrate the effect of different methods for handling medication use, we use data collected at baseline and consider three research questions:

i. The effect of systolic blood pressure (SBP) on the intima-media thickness (IMT), where the exposure is affected by medication.
ii. The effect of BMI on SBP, where the outcome is affected by medication.
iii. The effect of BMI on IMT, adjusted for SBP, where the confounder is affected by medication.

All methods described in Table 1 were applied to estimate the regression models corresponding to the three research questions stated above. The analyses were adjusted for potential confounders: BMI, sex, age, education level and smoking status.

In the Netherlands physicians prescribe blood pressure medication generally aiming at values below 140 mmHg (10). Therefore, we replaced treated SBP values by 150 mmHg in the substitution method [M4], and repeated it using 170 mmHg. For adding medication effect [M5], we followed previous literatures using the values 10 mmHg and 15 mmHg (4, 11). For regression calibration [M6], the assumed mean treatment effect was 15 mmHg; SD=10 mmHg.

For inverse probability weighting [M7], logistic regression was used to estimate the probability of medication use based on 21 covariates (see Supplementary Material 2 for details). The same covariates were used in the probit part of Heckman’s treatment model [M10] and in the multiple imputation approaches [M11-M13]. For quantile regression [M8], values of treated individuals were replaced by 150, 170 and 190 mmHg. For censored regression [M9] and imputation [M12], we used 140 mmHg and 160 mmHg as a clinical threshold for treatment prescription. For research question i) and iii) the outcome variable IMT was added to the imputation models (12). Ten imputed datasets were created in each imputation.

All analyses were performed using R version 3.6.1, with packages Survival v3.1-8 for (13) [M9], SampleSelection v1.2-6 (function treatreg) (14) for [M10], Quantreg v5.54 (15) for [M11], MICE v3.7.0 (16) with default options for [M11] and miceMNAR (17) for [M13]. R code for [M12] is provided in Supplementary Material 2.

Figure 1 presents effect estimates from the different methods for the three research questions. The results show that different methods can lead to quite different effect estimates in all three considered situations. This signals that choosing an appropriate method for handling measurements affected by medication use is essential for the validity of study results.

**Figure 1.**
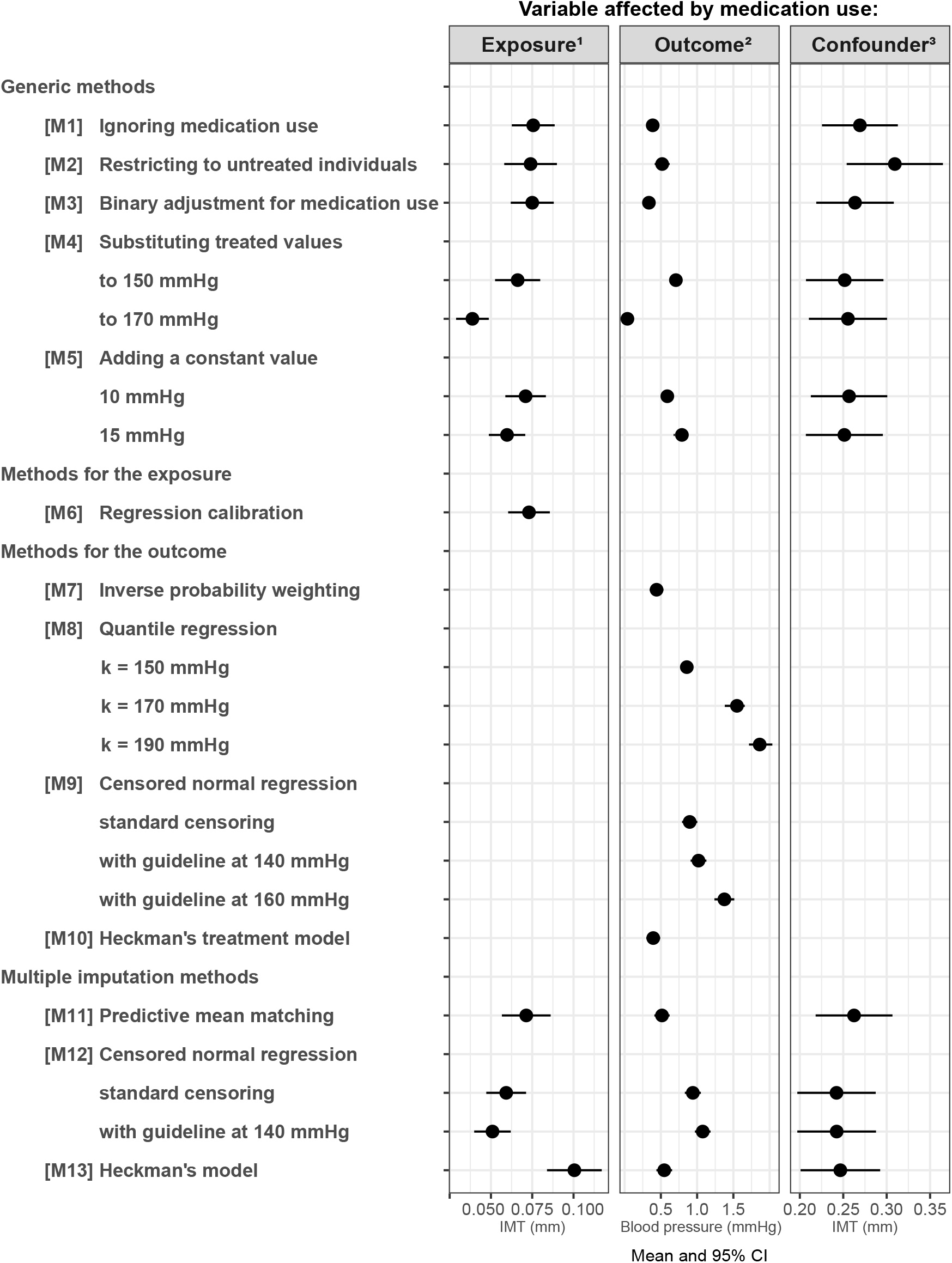
Regression coefficients and their 95% confidence interval estimated from the NEO data using the different methods to handle medication effect. ^1^Question 1: effect of SBP (mmHg) on IMT (mm), ^2^Question 2: effect of BMI (kg/m^2^) on SBP, Question 3: effect of BMI on IMT where SBP is one of the confounder. In all analyses, SBP was the variable affected by medication.

## 4. Simulation studies

To understand the results of the NEO study and provide recommendations, we performed several so-called real-life simulation studies. To mimic the NEO study as closely as possible, we used the baseline variables of the NEO participants (BMI, sex, age education and LDL cholesterol), but replaced SBP, antihypertensive medication prescription and IMT by simulated values which we let depend on the observed values of the other baseline variables. We generated different scenarios where blood pressure could be the exposure (scenario 1), the outcome (scenario 2) or the confounder (scenario 3). In each scenario we considered the research questions i), ii) and iii) of Section 3 respectively.

### 4.1 Simulation setting 1: Medication effect on the exposure

In this simulation setting, we are interested in the effect of SBP on IMT, with SBP affected by antihypertensive drug in some individuals. The *untreated SBP* depended linearly on *BMI, sex* (*man*=0, *women*=1), *age* and *education* (*low*=0, *high*=1), with parameter values closely corresponding to observed values in the NEO study:

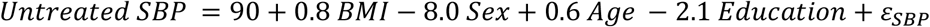

with the residual error *ε*_*SBP*_ normally distributed with mean 0 and SD 15.9 mmHg. The probability to receive medication depended on *BMI, sex, education* and the *untreated SBP* values:

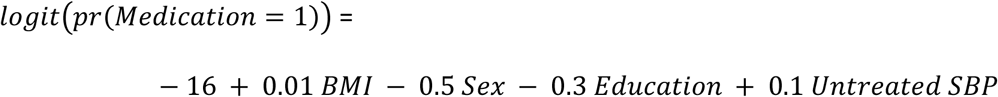

In this way, approximately 28% of the participants were treated for high SBP. For a SBP of 150 mmHg, the probability of receiving medication was approximately 11%, while for 180 mmHg the probability was 88%. The *Observed SBP* was lowered when medication was used:

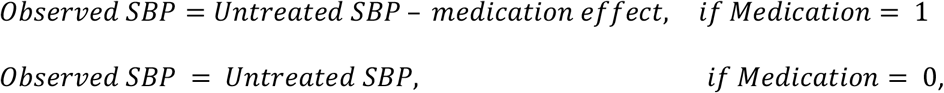

where the *medication effect* was generated from a normal distribution (30 mmHg, SD=10 mmHg). The outcome, IMT was generated as:

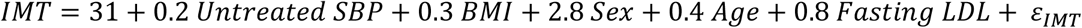

with *ε*_*IMT*_ following a normal distribution (0, SD= 9.2 mm). The relation between medication use and IMT is confounded by sex and BMI.

### 4.2 Simulation setting 2: Medication effect on the outcome

In Simulation setting 2, we consider the effect of BMI on untreated SBP. BMI was taken directly from the NEO data. Untreated SBP, medication prescription and the observed SBP were generated in the same way as in Simulation setting 1.

### 4.3 Simulation setting 3: Medication effect on a confounder

Here, we consider the effect of BMI on IMT measurement when adjusted for SBP. Untreated and observed SBP, medication prescription and IMT were generated as in Simulation setting 1.

### 4.4 Alternative simulation scenarios

Simulation setting 1, 2 and 3 were repeated while changing three parameters: i) The size of the mean treatment effect decreased from 30 mmHg to 10 mmHg. In this simulation 16% of the treated individuals’ SBP increased after medication. ii) The standard deviation of the treatment effect changed from 10 mmHg to 1 mmHg. iii) The percentage of individuals on medication increased from approximately 28% to 50% by changing the intercept of the logistic model for medication use.

### 4.5 Analysis

All methods [M1-M13] were applied to the simulated data sets in the same way as described in Section 3, except we used 20 mmHg and 30 mmHg to add to the treated SBP in [M5]. Analyses were adjusted for BMI, sex, age, education level and smoking status. Each simulation was repeated 1000 times. The estimates obtained when using the untreated SBP values were used as reference. Mean bias and mean squared error were calculated as overall measure of performance.

## 5. Results

### 5.1 Simulation setting 1: Medication effect on the exposure

Figure 2 (left) and Table 2 display the results of simulation setting 1. The results show that medication use cannot be ignored [M1]. Restricting the analysis to untreated individuals [M2] yielded estimates very close to the true values. In this setting medication use was affected by the exposure and several covariates, in which case one should <u>adjust</u> for all variables both affecting medication use and the outcome to prevent selection bias (18). Furthermore, there was no effect modification, meaning that the effect of SBP on the outcome in the subgroup of untreated individuals is the same as in the total population.

**Table 2.** Mean Coefficient, Standard Deviation, Bias and Mean Squared Error (MSE) for Three Main Simulation Settings.

| Methods | Exposure affected<sup>1</sup> |  |  |  | Variable affected by medication use:<br>Outcome affected<sup>2</sup> |  |  |  | Confounder affected<sup>3</sup> |  |  |  |
| --- | --- | --- | --- | --- | --- | --- | --- | --- | --- | --- | --- | --- |
|  | Mean | SD | Bias | MSEx1000 | Mean | SD | Bias | MSEx10 | Mean | SD | Bias | MSEx100 |
| **All untreated values known (true coefficient)** | 0.200 | 0.006 | 0.000 | 0.004 | 0.800 | 0.053 | 0.000 | 0.028 | 0.292 | 0.025 | 0.000 | 0.063 |
| **Generic methods** |  |  |  |  |  |  |  |  |  |  |  |  |
| [M1] Ignoring medication use | 0.165 | 0.007 | -0.035 | 0.127 | 0.483 | 0.048 | -0.317 | 1.028 | 0.372 | 0.025 | 0.080 | 0.703 |
| [M2] Restricting to untreated individuals | 0.200 | 0.008 | 0.000 | 0.006 | 0.564 | 0.054 | -0.236 | 0.586 | 0.295 | 0.030 | 0.003 | 0.091 |
| [M3] Binary adjustment for medication use | 0.182 | 0.007 | -0.018 | 0.037 | 0.533 | 0.048 | -0.267 | 0.736 | 0.302 | 0.025 | 0.010 | 0.073 |
| [M4] Substituting treated values |  |  |  |  |  |  |  |  |  |  |  |  |
| to 150 mmHg | 0.223 | 0.007 | 0.023 | 0.058 | 0.532 | 0.040 | -0.268 | 0.734 | 0.333 | 0.026 | 0.041 | 0.236 |
| to 170 mmHg | 0.180 | 0.006 | -0.020 | 0.044 | 0.743 | 0.052 | -0.057 | 0.060 | 0.318 | 0.025 | 0.026 | 0.130 |
| [M5] Adding a constant value |  |  |  |  |  |  |  |  |  |  |  |  |
| 20 mmHg | 0.200 | 0.006 | 0.000 | 0.004 | 0.694 | 0.051 | -0.106 | 0.138 | 0.313 | 0.025 | 0.021 | 0.107 |
| 30 mmHg (true value) | 0.187 | 0.006 | -0.013 | 0.021 | 0.800 | 0.056 | 0.000 | 0.031 | 0.302 | 0.025 | 0.010 | 0.073 |
| **Methods for exposure affected** |  |  |  |  |  |  |  |  |  |  |  |  |
| [M6] Regression calibration | 0.199 | 0.006 | -0.001 | 0.004 | - | - | - | - | - | - | - | - |
| **Methods for outcome affected** |  |  |  |  |  |  |  |  |  |  |  |  |
| [M7] Inverse probability weighting | - | - | - | - | 0.521 | 0.056 | -0.279 | 0.810 | - | - | - | - |
| [M8] Quantile regression |  |  |  |  |  |  |  |  |  |  |  |  |
| k = 150 mmHg | - | - | - | - | 0.429 | 0.037 | -0.371 | 1.390 | - | - | - | - |
| k = 170 mmHg | - | - | - | - | 0.948 | 0.070 | 0.148 | 0.268 | - | - | - | - |
| k = 190 mmHg | - | - | - | - | 1.052 | 0.100 | 0.252 | 0.735 | - | - | - | - |
| [M9] Censored normal regression |  |  |  |  |  |  |  |  |  |  |  |  |
| standard censoring | - | - | - | - | 0.749 | 0.054 | -0.051 | 0.055 | - | - | - | - |
| with guideline at 140 mmHg | - | - | - | - | 0.756 | 0.054 | -0.044 | 0.049 | - | - | - | - |
| with guideline at 160 mmHg | - | - | - | - | 0.879 | 0.063 | 0.079 | 0.102 | - | - | - | - |
| [M10] Heckman's treatment model | - | - | - | - | 0.660 | 0.083 | -0.140 | 0.265 | - | - | - | - |
| **Multiple imputation methods** |  |  |  |  |  |  |  |  |  |  |  |  |
| [M11] Predictive mean matching | 0.208 | 0.008 | 0.008 | 0.013 | 0.555 | 0.056 | -0.245 | 0.632 | 0.332 | 0.026 | 0.040 | 0.228 |
| [M12] Censored normal regression |  |  |  |  |  |  |  |  |  |  |  |  |
| standard censoring | 0.221 | 0.007 | 0.021 | 0.049 | 0.750 | 0.055 | -0.050 | 0.055 | 0.286 | 0.025 | -0.006 | 0.066 |
| with guideline at 140 mmHg | 0.212 | 0.007 | 0.012 | 0.019 | 0.756 | 0.055 | -0.044 | 0.050 | 0.291 | 0.025 | -0.001 | 0.063 |
| [M13] Heckman's model | 0.182 | 0.008 | -0.018 | 0.039 | 0.737 | 0.127 | -0.063 | 0.201 | 0.311 | 0.030 | 0.019 | 0.126 |

<sup>1</sup>Scenario 1: Effect of systolic blood pressure on IMT measurement. <sup>2</sup> Scenario 2: Effect of BMI on systolic blood pressure. <sup>3</sup> Scenario 3: Effect of BMI on IMT measurement, where systolic blood pressure is one of the confounders. In all scenarios, systolic blood pressure was the variable affected by medication.

**Figure 2.**
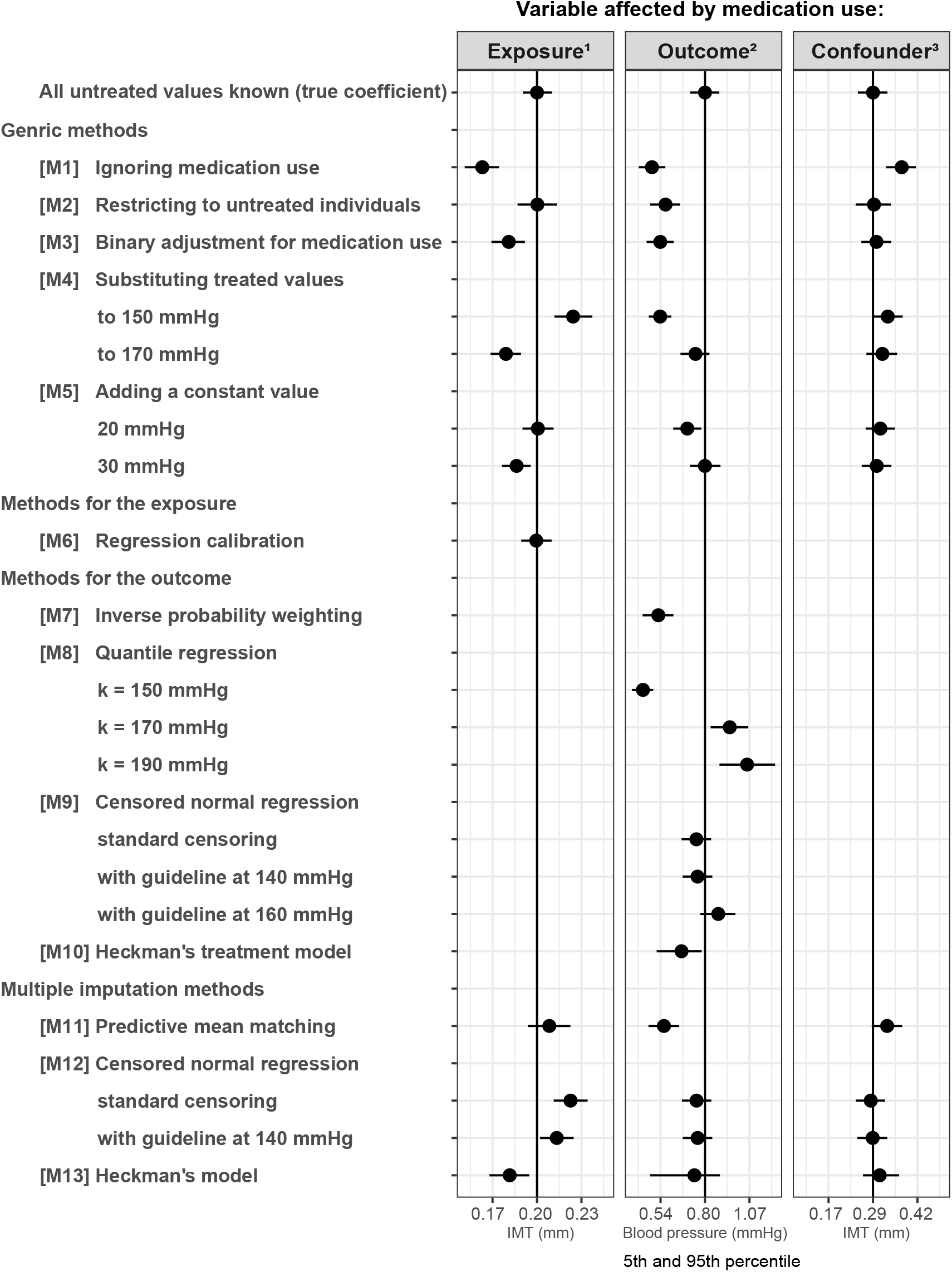
Regression coefficients and their 5th and 95th percentile estimated from simulation setting 1, 2 and 3. Results are standardized based on the mean and standard deviation of the true coefficients in each simulation setting. One grid unit represents 2.5 standard deviation. Simulation 1: effect of SBP on IMT, Simulation 2: effect of BMI on SBP, Simulation 3: effect of BMI IMT, where SBP is one of the confounders. In all scenarios, SBP was the variable affected by medication use.

Binary adjustment for medication use [M3] did not work well. In our simulation, medication effect was generated with a large variability. This random variability in medication effect attenuated the association between SBP and IMT in the *treated* individuals and led to bias toward the null in the overall effect. The method worked better when the variance of the medication effect was smaller (Supplementary material 3). Substituting treated values [M4] did not perform well in any of the scenarios. The method cannot reconstruct the original distribution of the exposure and therefore in general will yield biased results.

Adding 30 mmHg [M5], which was the true mean medication effect in our simulations did not perform well either. The reason is that the medication effect was generated with SD=10 mmHg. Therefore, by adding 30 mmHg to all treated SBP values, we reconstruct untreated SBP with random measurement error. Random measurement error in exposures will bias the estimates in a regression model (14). The method performed better when the random variation of the medication effect was smaller (Supplementary material 3). Regression calibration [M6] yielded unbiased results in all our simulations scenarios, assuming that true medication effect and standard deviation are known.

None of the multiple imputation methods [M11-M13] yielded valid results. A possible explanation is that the imputation models included the outcome which does not corresponds to how medication use was generated in our simulations.

### 5.2 Simulation setting 2: Medication effect on the outcome

Figure 2 (middle) and Table 2 show the results of Simulation setting 2. Ignoring mediation use [M1], restricting to untreated subgroup [M2] and binary adjustment for medication use [M3] yielded biased results. As the outcome determines medication use directly, adjusting or selecting based on medication use [M2 & M3] implies selection based on outcome values, and this will in general lead to selection bias (19, 20).

Substituting method [M4] using 150 mmHg led to a large underestimation. It performed better when 170 mmHg was used, which is slightly higher than the mean untreated SBP in the treated individuals (164 mmHg). Regardless of the substituting values, however, the method cannot reconstruct the original distribution of the outcome

Adding 30 mmHg [M5] yielded unbiased results in all simulation settings (Supplementary Material 4). Unlike in Simulation setting 1, adding the true mean medication effect yields valid results irrespectively of the amount of variance in the medication effect.

Inverse probability weighting [M7] resulted in large bias. Quantile regression [M8], for all chosen replacement values performed poorly. In our simulation setting, more than 50% were using antihypertensive drug among the individuals with very high BMI. Therefore, the median SBP conditional on high BMI was affected by whichever value used for substitution.

Censored normal regression [M9] performed reasonably well when the simple censoring method was used or when clinical guideline set to 140 mmHg was applied. However, in alternative scenarios with a smaller medication effect, the results were off (Supplementary Material 4). One reason is that in these scenarios the treated SBP was sometimes higher than the untreated SBP. This violates the assumption that untreated values are at least as high as untreated values (1). Heckman’s treatment model [M10] performed less well in our main scenario, which contrasts the results reported by Spieker et al. (2, 7). This model assumes that the residual variances of two linear regression models, one for untreated individuals and the other for treated individuals, are equal. This assumption was violated in our main simulation scenario, as we simulated a medication effect with a large random variability. This reflects the reported instability of Heckman’s treatment (21, 22). In the scenarios with a smaller variance in the medication effect, Heckman’s treatment model outperformed the censored regression (Supplementary Material 4).

Multiple imputation with predictive mean matching [M11] resulted in bias. Results of multiple imputation with censored regression [M12] were only slightly biased, but for smaller medication effect the method performed less well. Multiple imputation with Heckman’s model [M13] yielded in some cases a large underestimation of the effect in all scenarios.

### 5.3 Simulation setting 3: Medication effect in a confounder

Figure 2 (right) and Table 2 show the results of Simulation setting 3. Ignoring the medication effect [M1] resulted in bias. Restricting to untreated individuals [M2], which is the same as adjusting for confounding by restriction, performed well. The method will yield valid estimation under the conditions as in Simulation setting 1, that is with proper adjustment for variables affecting both medication use and the outcome. Binary adjustment for medication use [M3] yielded results close to the truth. Substitution methods [M4] were biased. As the distribution of untreated SBP could not be correctly reconstructed, its effect was not correctly adjusted for.

Adding 30 mmHg [M5] yielded a very small upward bias. This is due to the random measurement error introduced by the method. It has been known random measurement error in exposures attenuates the effect, while random measurement in confounders can lead to overestimation (23, 24).

Multiple imputation with censored regression [M12] yielded results close to the truth, especially when clinical guideline information was incorporated, and performed better than multiple imputation with Heckman’s model [M13]. All results were consistent in the alternative simulation scenarios (Supplementary Material 5).

## 6. Guidance on how to optimally handle measurements affected by medication use

When interest is in the relation between the unaffected variables, ignoring medication use will in general yield biased results regardless whether the exposure, outcome or confounder is affected by medication. To obtain valid estimates, methods for handling medication use are needed. In this section, we provide guidance on how to handle an exposure, an outcome or a confounder affected by medication use.

### What to do when the exposure is affected by medication?

- Performing analysis on the untreated individuals [M2] is a valid approach and will not lead to a large loss in power if the number of treated individuals is relatively low. However, there are two things to consider when applying this method: i) One should adjust for variables which both affect medication use and the outcome. ii) The result cannot be generalized to the total population if the effect of the exposure on the outcome is heterogenous.
- Regression calibration [M6] may be used, but requires an external estimate of the medication effect with its standard deviation.

### What to do when the outcome is affected by medication?

- When an estimate of the mean medication effect is available, it could be added to the measurements of treated individuals. This method was also advocated by Tobin et al. (1). Like them, we also highly recommend to perform sensitivity analysis with several different values to determine the stability of effect estimates.
- Quantile (median) regression [M8] can be used when less than 50% of the individuals are treated at any value of the exposure. The method does not require knowledge on the medication effect and can yield robust estimates but with lower power (6) than other methods.
- The advantage of censored normal regression [M9] or multiple imputation with censored normal regression [M12] is that no treatment effect needs to be specified. However, the method assumes that the observed values are lower than the untreated values, which could be violated when the treatment is ineffective. Furthermore, the method assumes non-informative censoring which is likely to be violated in most clinical settings. In our simulation, we relaxed this assumption by incorporating knowledge from a clinical guideline into a censoring mechanism. Both in study of Tobin et al. and in our main simulation study, the method was rather robust against the violation of non-informative censoring assumption.
- Heckman’s treatment model works well only if the treatment effect has small variance.

### What to do when the confounder is affected by medication?

- Restricting the analysis to untreated individuals [M2] is a valid approach, with the same considerations as for the exposure affected by treatment.
- Using a binary indicator [M3] is a reasonable solution.
- Adding the true mean medication effect to the treated individuals [M5] performs relatively well.

## 7. Discussion

Our simulation study showed that the problem of variables affected by medication use should not be ignored, and proper methods are needed to avoid potential bias. Different methods are needed depending on whether the exposure, the outcome or a confounder is affected by medication. Additional information, such as medication prescription patterns in clinical settings and presence of effect heterogeneity should also be considered carefully. Accordingly, all methods need to be used with caution.

One important consideration is the trade-off between the robustness of a method and availability of external information. Methods that use external information on the medication effect, such as adding the mean medication effect or regression calibration, performed well when the external information was correct. However, such information is not always available. Other methods such as censored regression, Heckman’s treatment model or multiple imputation methods, do not require assumptions on the medication effect. However, they rely on other assumptions and can perform suboptimal if they are violated.

We aimed our simulation scenarios to resemble realistic clinical situations, instead of creating an ideal scenario for a particular method. It is likely that assumptions required for statistical methods will not all be met in clinical data. Therefore, it is relevant to know which methods are robust against violation of assumptions. We encourage researchers to perform more often real-life simulations as we did when generating simulations based on the NEO study data.

One limitation of our study is that we did not considered situations where more than one variable is affected by medication. Additionally, our study focused on the methods applicable for cross-sectional analyses. Other approaches may be available when there is an interaction by medication (25), when effect modifiers are associated with medication use (7), in longitudinal settings (26) or in the presence of interaction or mediation by time-varying treatment (27). Furthermore, we focussed on linear regression models, but our recommendations for exposures and confounders will also hold for regression models with a binary or survival outcome.

In summary, the optimal strategy for handling measurements affected by medication depends on whether the medication effect is on the exposure, the outcome or a confounder. When deciding which strategy to use, we urge researchers to critically consider the processes of medication prescription and what information on medication effects are available.

## Data Availability

The simulated data that support the findings of this study are available from the corresponding author upon reasonable request. The Netherlands Epidemiology of Obesity (NEO) data are not publicly available due to privacy or ethical restrictions.

## Notes

### Competing Interest Statement

The authors have declared no competing interest.

### Funding Statement

This study did not receive any funding

### Author Declarations

The medical ethical committee of the Leiden University Medical Center gave ethical approval for the NEO study. The use of the NEO study data was approved by the NEO study board.

