## Supplementary Material 1 for "A comparison of different methods for handling measurements affected by medication use"

**Supplementary Material 1: Detailed description on methods for handling medication effect**

We consider the situation where for some individuals a variable is affected by medication use (e.g., blood pressure affected by antihypertensive drug), while the linear relationship between variables when no one is affected by medication is of interest. For convenience, we assume that medication is taken when values are high, aiming to lower the values. Depending on the research question, the variable(s) affected by medication use can be the exposure, a confounder or the outcome in an analysis. The different methods to handle medication use are :

### NAÏVE METHODS

#### *M1. Ignoring medication use*

Measurements affected by medication are used in the analysis as they are observed.

#### *M2. Selecting untreated individuals*

Only the individuals who are not receiving medication are included in the analysis.

#### *M3. Adjusting for medication use by adding a binary indicator variable to the regression model*

An indicator for medication use is added as a covariate in the regression model.

#### *M4. Substituting measurements of treated individuals with a fixed value*

Suggested by Hunt et al (1), measurements affected by medication can be substituted with a pre-specified value. For example, when guidelines indicate that blood lowering medication should be prescribed for blood pressures over 140 mmHg, a value higher than 140 mmHg can be used as a substitution.

#### *M5. Adding a constant value to observations of treated individuals*

When the effect of medication on the variable of interest is approximately known, the mean treatment effect can be added to the observed measurements of treated individuals (2, 3). For blood pressure, for example, some authors added 10 mmHg to the systolic blood pressure and 5 mmHg to the diastolic blood pressure when individuals are using antihypertensive medication (4, 5). These values were based on known average treatment effects from a clinical trial (6). However, this is not a set rule and could be adapted

### METHODS FOR A MEDICATION EFFECT IN EXPOSURE

#### *M6. Regression calibration*

There is a vast amount of literature to deal with measurement error in the covariates of a regression model (7, 8). A simple method is regression calibration, where the expected untreated values replace the measurements affected by medication. The expected untreated values are estimated by the observed values and other covariates. The method needs an educated guess of the mean *and* standard deviation of the medication effect. These may be obtained from previous clinical trials or observational studies where the effect of treatment is studied.

For individuals on medication their observed measurement *X* is replaced by $\lambda(X - \bar{X}) + \bar{X}$ + *mean medication effect*; with $\bar{X}$, the mean value of *X* for those using medication and λ, so-called reliability ratio (9). Reliability ratio is equal to $\lambda = 1 - SD(med)^2/SD(X|Z)^2$; with *SD(med)*, the standard deviation of the medication effect and *SD(X|Z)*, the standard deviation of *X* for the medication users adjusted for *Z*, a set of other covariates in the regression model.

**METHODS FOR A MEDICATION EFFECT IN THE OUTCOME**

***M7.*** **Inverse probability weighting (Sampling weights)**

In this approach, treated individuals are removed in the analysis, and more weight is given to individuals who are untreated but have a similar profile as treated individuals (10, 11). First, the probability of receiving medication for each individual is estimated by logistic regression. Then the untreated individuals are weighted by 1/(1-probability to receive medication). This creates a pseudo population with the same characteristics as the original population but where no one is treated.

***M8. Quantile regression***

White et al. (12) proposed to use quantile regression for outcomes affected by medication use. In this approach, the median outcome is modeled as a function of covariates. The method assumes the untreated values would have been above the median conditional on covariates for individuals on medication. The treated individuals' outcome values are replaced by *k,* that is, any value higher than the conditional median, after which a median regression model can be fitted.

***M9. Censored normal regression.***

An alternative approach is to use methods for censored outcomes (2, 3), such as censored normal regression, which assumes a normal underlying distribution of the untreated outcome. This method is also known as tobit regression. Measurements of treated individuals are considered to be censored observations, where the untreated values are assumed to be at least as high as the observed values affected by treatment. An advantage of this method is that no assumptions on the treatment effect size are needed. However, non-informative censoring is assumed. The non-informative censoring implies that conditional on covariates, the probability of receiving treatment does not depend on the untreated values. This assumption is likely to be invalid, as individuals with higher values are more likely to be treated. Previous simulations showed good performance in realistic scenarios (2). However, recent literature showed that the method performed poorly under certain scenarios (13).

More complex censoring mechanisms can also be used to resemble realistic clinical settings. For example, when a clinical guideline suggests starting treatment for values above a certain threshold δ, this information can be incorporated. In this case, the untreated values are assumed to be higher than the observed measurements *and* higher than the threshold. That is, for the treated observations, we assume that:

$$\begin{cases} Untreated\ value\ \geq \delta, & if\ observed\ value < \delta \\ Untreated\ value\ \geq observed\ value, & if\ observed\ value \geq \delta \end{cases}$$

The threshold value of $\delta$ is obtained using external knowledge of the clinical setting.

***M10. Heckman's treatment effects model***

Heckman's treatment effects model originates from economics and can account for non-random sample selection (13-15). Spieker et al. (13, 15) used this model for handling outcomes affected by medication use. This model assumes that treatment assignment depends on the untreated values where higher values are more likely to be treated and treatment results in a "structural shift" of the mean outcome. In the standard treatment effect model, this treatment effect does not depend on covariates (13), but it is possible to extend this model to incorporate effect modification (15).

Technically, the method assumes that there is an unobserved latent variable that determines treatment. If its value is above 0, treatment is prescribed. The latent variable is correlated with the original untreated values, so people with higher untreated values are more likely to be treated. Parameters are estimated by joint modeling of i) a linear regression model for the effect of exposure on the untreated blood pressure, ii) the same linear regression model for the effect of exposure on the treated blood pressure, with a lower constant term which reflects the effect of treatment and iii) a probit model for the probability of medication prescription (13, 15). Both the linear regression model and the probit model may depend on other covariates.

**MULTIPLE IMPUTATION APPROACHES**

Untreated values of individuals on treatment can be considered missing, and multiple imputation methods can be used to handle these missing values. The method can be applied in many different ways under different assumptions. We considered three multiple imputation approaches that are based on various assumptions.

***M11. Multiple imputation with predictive mean matching via a linear regression model***

For a numerical variable with missing values, the default multiple imputation option is chained equation with predictive mean matching via a linear regression model with main effects of the covariates. This imputation method is readily available in many standard statistical software packages. Note that the method assumes that the data is missing at random.

***M12. Multiple imputation with censored normal regression***

Instead of using linear regression as imputation model, censored normal regression may be used to predict missing values (16). This may be done under the different censoring mechanisms we discussed for [M9]. While regular censored normal regression can only be used when medication effect is on the outcome, multiple imputation with censored normal regression does not have this restriction.

***M13. Multiple imputation with Heckman's model***

Galimard et al. developed an imputation approach for missing not at random data using a Heckman's model (17). Again, this multiple imputation approach can be used both when the outcome is affected by medication and when exposures or confounders are affected.
