## Supplementary Material 2 for "A comparison of different methods for handling measurements affected by medication use"

**Supplementary Material 2: Detailed application of the methods**

**1) Covariates used for inverse probability weighting [M7] and the imputation methods [M11-M13]**

Sex, age, BMI, total body fat, waist circumference, hip circumference, education level, income, smoking status, ethnicity, alcohol intake, total amount of leisure, glucose, insulin, glycated hemoglobulin A1C, triglycerides, HDL cholesterol, LDL cholesterol and medication use for glucose, lipid and depression.

**2) R syntax for censored normal imputation [M12]**

```
# Function to draw from a truncated normal distribution, range lwb-upb
rnorm.trunc <- function(n,mean,sd, low=-Inf, upp=Inf)
{U <- runif(n,0,1)
qnorm(pnorm(low, mean = mean, sd = sd)+
    (pnorm(upp, mean = mean, sd = sd)-pnorm(low, mean = mean, sd = sd))*U, mean = mean, sd =
sd)
}

# impute censored normal
mice.impute.censnorm <-
  function (y, ry, x, wy = NULL,ycens, ...)
  {
    #1 prepare data
    wy <- !ry # wy= TRUE indicates that value should be imputed
    x <-  as.matrix(x)
    m <- ncol(x)+1

    # 2. estimate coefficients censored model
    fit <- survreg(Surv(ycens, ry) ~ x, dist='gaussian')
    beta <- coefficients(fit)
    sigma <- fit$scale
    #    print(fit)

    #3. generate new beta and sigma for bayesian drawings
    df <- max(length(y[ry]) - ncol(x), 1)
```

```
  rv <- t(chol((vcov(fit)[1:m,1:m])))
  beta.star <- beta + rv %*% rnorm(ncol(rv))
  sigma.star <- sqrt(df*sigma^2/rchisq(1, df))

  #4. Draw new observations
  mean.star <- cbind(1,x[wy, , drop = FALSE]) %*% beta.star
  vec<- rnorm.trunc(nrow(mean.star),mean.star,sd=sigma.star, low=ycens[wy])
  return(vec)
}
```
