## Supplementary Material 4 for "A comparison of different methods for handling measurements affected by medication use"

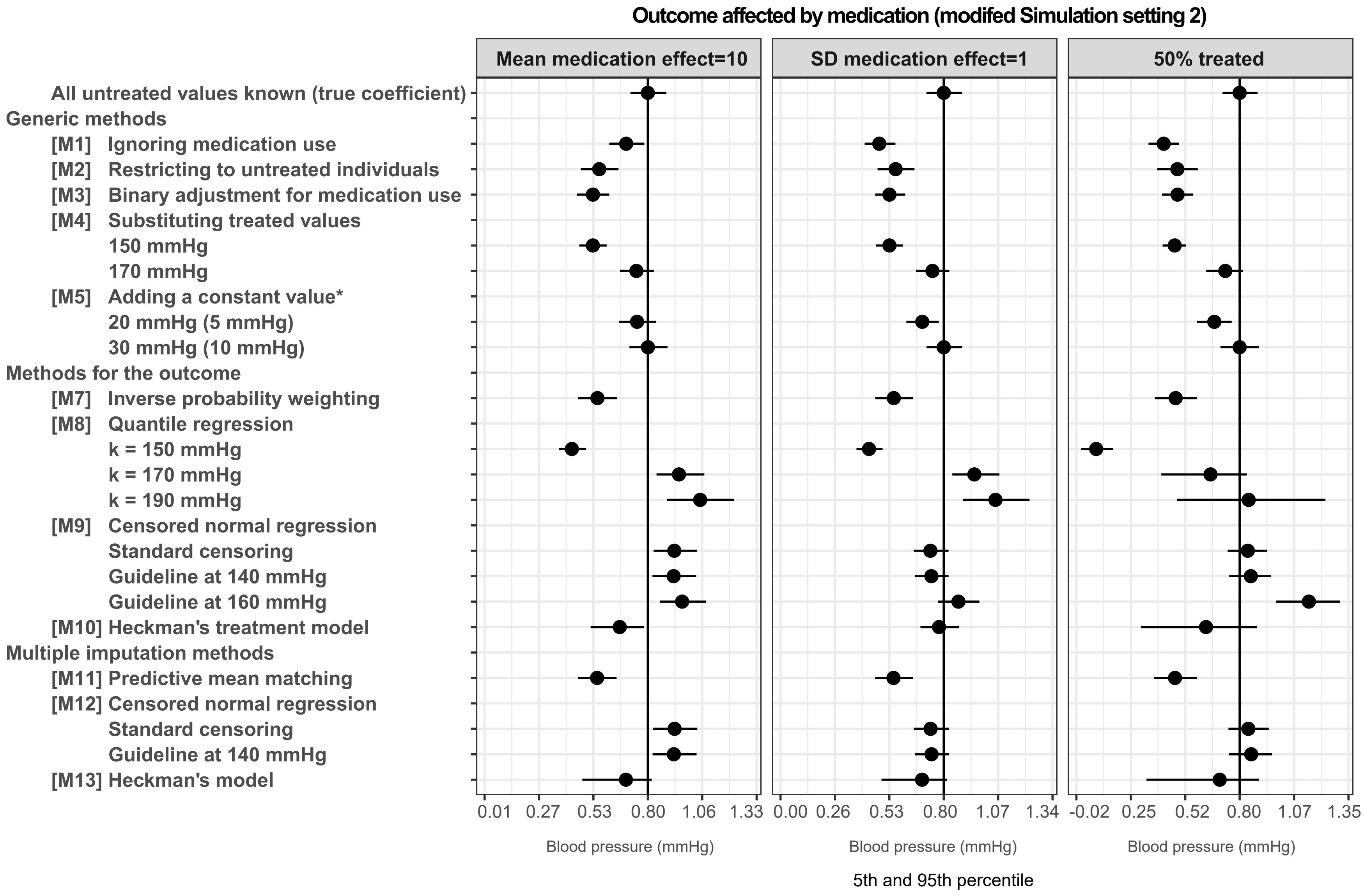


**Supplementary Material 4.** Regression coefficients and 5th and 95th percentile estimated from modified scenarios of Simulation setting 2, where we estimated the effect of BMI on SBP. One grid unit represents 2.5 standard deviation of the true coefficient, estimated from 1000 simulation runs.
*In the scenario where mean medication effect was 10 mmHg, 5 mmHg and 10 mmHg were added.
